# The COVID-19 outbreak in Sichuan, China: epidemiology and impact of interventions

**DOI:** 10.1101/2020.07.20.20157602

**Authors:** Quan-Hui Liu, Ana I. Bento, Kexin Yang, Hang Zhang, Xiaohan Yang, Stefano Merler, Alessandro Vespignani, Jiancheng Lv, Hongjie Yu, Wei Zhang, Tao Zhou, Marco Ajelli

**Affiliations:** College of Computer Science, Sichuan University, Chengdu, China; Department of Epidemiology and Biostatistics, School of Public Health, Indiana University Bloomington, IN, USA; Department of Engineering and Computer Science, New York University Shanghai, Shanghai, China; Institution of New Economic Development, Chengdu, China; Bruno Kessler Foundation, Trento, Italy; Laboratory for the Modeling of Biological and Socio-technical Systems, Northeastern University, Boston, MA USA; ISI Foundation, Turin, Italy; School of Public Health, Fudan University, Key Laboratory of Public Health Safety, Ministry of Education, Shanghai, China; West China Biomedical Big Data Center, West China Hospital, Sichuan University, Chengdu, China; Big Data Research Center, University of Electronic Science and Technology of China, Chengdu, China

**Keywords:** COVID-19, SARS-CoV-2, Interventions, Averted cases, Reproduction number

## Abstract

In January 2020, a COVID-19 outbreak was detected in Sichuan Province of China. The aim of this work is to characterize the epidemiology of the Sichuan outbreak and estimate the impact of the performed interventions. We analyzed patient records for all laboratory-confirmed cases reported in the province for the period of January 21 to March 16, 2020. To estimate the basic and daily reproduction numbers, we used a Bayesian framework. In addition, we estimate the number of cases averted by the implemented control strategies. The outbreak resulted in 539 confirmed cases, lasted less than two months, and no further local transmission was detected after February 27. The median age of local cases was 8 years older than that of imported cases. Severity of symptoms increased with age. We estimated R_0_ at 2.4 (95% CI: 1.6–3.7). The epidemic was self-sustained for about 3 weeks before going below the epidemic threshold 3 days after the declaration of a public health emergency by Sichuan authorities. Our findings indicate that, were the control measures be adopted four weeks later, the epidemic could have lasted 49 days longer (95%CI: 31-68 days), causing 9,216 (95%CI: 1,317-25,545) more cases and possibly overwhelming Sichuan healthcare system.

## 1. Background

SARS-CoV-2 has been incredibly successful in spreading swiftly from Wuhan, Hubei Province in China. Cases numbers rocketed and the disease spread through China, quickly entering an exponential growth phase [1]. Non-pharmaceutical interventions were quickly put in place both at the epicenter of the outbreak and country-wide, culminating in the implementation of the lockdown of entire populations with the goal to suppress or mitigate the epidemic to prevent overwhelming health care systems [2,3].

Chinese provinces outside Hubei represent an important example of successful local containment of COVID-19 outbreaks and thus represent a valuable source of information for other countries as well. Sichuan is one of the largest provinces of China with a population size of about 83-million individuals and a major transport hub in Southwestern China. Sichuan was one of the first provinces outside Hubei to record COVID-19 cases with a first importation from Wuhan detected on January 21, 2020 [4]. Then, despite the importations of several tens of cases from Wuhan/Hubei over a relatively short period of time (less than 2 months), a major epidemic wave was avoided. As of June 22, 2020, 589 cases are reported by the CDC of Sichuan province [5].

The aim of this study is to describe the epidemiological characteristics of the COVID-19 outbreak in the Sichuan and to shed light on its successful local containment. By using a Bayesian approach based on the renewal equation [6,7], we estimated the basic and daily reproduction numbers. The posterior distribution of the reproduction number has been used to project the number of cases in case the adopted interventions had started between one and four weeks later. This allowed us to estimate the number of cases, including severe and critical ones, which were averted by the timely implemented control strategies and population awareness.

## 2. Methods

### (a) Data

#### Case definition

Case definitions and the types (paucisymptomatic, symptomatic, severe, and critical) classified are according to the fifth version of “Guideline on diagnosis and treatment of novel coronavirus infected pneumonia (NICP)” issued by China CDC on February 4, 2020 [5]. Briefly, mild cases are defined as cases showing mild clinical symptoms and no radiographic evidence of pneumonia. Symptomatic cases present clinical symptoms, such as fever and respiratory symptoms as well as radiographic evidence of pneumonia. Severe cases have to meet one of the following conditions: i) respiratory rate interval ≥30 b.p.m.; ii) SpO_2_ (saturation of peripheral oxygen) ≤93% at rest; iii) PaO2/FiO2 ≤ 300mmHg (1mmHg=0.133kPa). Critical cases have to meet one of the following conditions: i) respiratory failure and consequent needs of mechanical ventilation; ii) shock; iii) require intensive care because of multiple organ dysfunction.

#### Case data

Patient records were provided by Sichuan CDC [4], which curates a centralized database of hospitals records including all hospitals in each prefecture of Sichuan. The dataset contains information on age, sex, location of detection, exposure history, dates of symptom onset, hospital admission, and official reporting to define demographic characteristics of cases and estimate main time-to-event intervals and the daily reproduction number. Information on the travel history in the last 14 days, the clinical types, and the reporting prefecture were also recorded.

#### Performed interventions

All schools of Sichuan Province were closed on January 18, 2020. Sichuan declared the top-level public health emergency and established an emergency command center on January 24, 2020. Following this, gathering activities and entertainment (e.g., sports events) were suspended and public libraries closed. On January 26, the Sichuan government announced a strict set of measures to deal with the outbreak, including case isolation, tracing and screening of contacts of confirmed cases, quarantine of travelers from affected areas, and screening of people’s temperature in public places. As of February 25, 2020, fourteen prefectures in Sichuan had no new confirmed cases for a week and the government decided to allocate provincial resources to deal with the epidemic in high-risk areas, while gradually starting to relax the interventions in medium and low-risk areas.

### (b) Statistical and modeling analysis

#### Descriptive statistics

We used the patient records to calculate the age distribution and gender of cases disaggregated by case severity. Further, we distinguished between locally acquired infections and those with travel history from Wuhan/Hubei. Additionally, we calculated distributions of time intervals from symptom onset to hospital admission and from symptom onset to reporting for those cases where all information was available.

#### Reproduction number

The basic reproduction number R_0_ represents the mean number of secondary cases generated by a primary infector during the exponential growth phase of the epidemic, before interventions are applied and when the depletion of susceptible individuals is negligible [8]. The daily reproduction number R(t) represents the mean number of secondary cases generated by a primary infector at time *t* [9]. The daily reproduction number is useful to track the effectiveness of performed control measures, which aims to push it below the epidemic threshold (corresponding to R(t)=1). Moreover, R(t) accounts for other factors affecting the spread of the epidemic such as the behavioral response of the population over time and the depletion of susceptible individuals in the population.

To estimate R(t), we use the same methodology adopted by Zhang et al. [10], which was adjusted from [6,7] to distinguish between locally acquired and imported cases. Briefly, we assume that the daily number of new cases (date of symptom onset) with locally acquired infection *L(t)* can be approximated by a Poisson distribution according to the renewal equation

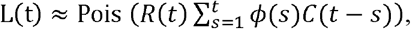

where *C(t)* is the number of new cases (either locally acquired or imported) at time t (date of symptom onset), R(t) is the effective reproduction number at time *t* and *ϕ* is the generation time distribution. To estimate the time between consecutive generations of cases, we adopted the serial interval (which measures the time difference between the symptom onset of the infectors and of her/his infectees) estimated from the analysis of the first few clusters of COVID-19 cases detected in Wuhan before the implementation of the interventions, namely a gamma distribution with mean 7.5 days (shape=4.87, rate=0.65) [11].

The likelihood Λ of the observed time series cases from day 1 to T can be written as

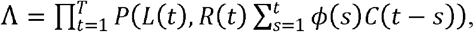

where *P(x,y)* is the Poisson density distribution of observing × events, given the parameter y.

We then use Metropolis-Hastings MCMC sampling to estimate the posterior distribution of R(t). The Markov chains were run for 100,000 iterations, considering a burn-in period of 10,000 steps, and assuming non-informative prior distributions of R(t) (flat distribution in the range (0-1000]). Convergence was checked by visual inspection by running multiple chains starting from different starting points.

To estimate R_0_, we estimated a constant daily reproduction number over a time window early on in the outbreak and before the implementation of interventions [7]. Specifically, we estimated R_0_ over the 1-week time window before the declaration of the outbreak, namely from January 18 to 24 and, when the outbreak was growing exponentially. In addition, as a sensitivity analysis, we estimated R_0_ over a more conservative 2-week time window (from January 11 to 24, 2020).

#### Counterfactual scenarios

To estimate the number of cases averted by the policies implemented in Sichuan since the declaration of a public health emergency, we provided a set of counterfactual scenarios where we consider different starting dates of the interventions. To project the number of new COVID-19 cases (assuming a different starting date of the interventions), we use the renewal equation [7], which we already used to estimate the daily reproduction number. The projected number of new cases at time t can thus be estimated as:

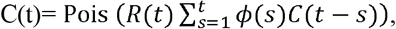

where the notation is the same used in the previous section.

The proposed counterfactual scenarios consider a schematic representation of the dynamics of R(t). In particular, to mimic the estimated dynamics of R(t) (see Section Results) we assume R(t)=R_0_ before the implementation of the control strategies; then we consider R(t) to linearly decrease over a 1-week time window to its final constant value (R_final_) estimated over the period from February 1 to the end of the epidemic.

Four counterfactual scenarios are considered, each one accounting for a different starting date of the interventions ranging from 1 week (January 31) to 4 weeks (February 21) after the actual declaration of the public health emergency from Sichuan health authorities on January 24. Each counterfactual scenario is based on 1,000 simulations, each one considering a value of R_0_ and a value of R_final_, sampled from the two estimated posterior distributions. It is important to stress that over the projection periods, the depletion of the susceptible population is negligible as compared with actual size of the Sichuan population (about 83,000,000 individuals [12]).

In addition to the main analysis described above, we consider two sensitivity analyses. In the first one, we consider a lower value of R_0_, as estimated over a 2-week time period before the declaration of the public health emergency. In the second one, instead of a linear decrease from R_0_ to R_final_, we consider an instantaneous switch between the two values occurring on the day when the public health emergency is declared.

#### Averted cases

We define the number of averted cases as the difference between the final number of cases projected by the model and the actual number of reported cases. Similarly, we defined the number of averted severe and critical cases by multiplying the projected number of cases by the probability of developing severe or critical condition as estimated from the analysis of the patient records.

### (c) Ethics approval

The study was approved by the Clinical Trials and Biomedical Ethics Committee of West China Hospital, Sichuan University (No. 2020190).

## 3. Results

### Outbreak description

As of the March 16, 2020 a total of 539 cases were confirmed in Sichuan, including four asymptomatic subjects, 115 mild cases, 331 symptomatic cases, 57 severe cases, and 32 critical cases that required ICU treatment (Table 1). Among these confirmed cases, 253 had travel history from/to Wuhan/Hubei, 200 cases were locally acquired, and for the remaining 86 cases not known (Tab. 1 and Fig. 1a). Thus, the Sichuan outbreak was characterized by a combination of local transmission and case importations.

**Table 1.** Characteristics of laboratory-confirmed COVID-19 cases in Sichuan disaggregated by case severity.

| Characteristics/Case | Total | Mild | Symptomatic | Severe | Critical |
| --- | --- | --- | --- | --- | --- |
| Male - no./total no. (%) | 285/539 (52.9%) | 56/115 (48.7%) | 177/331 (53.5%) | 32/57 (56.1%) | 17/32 (53.1%) |
| Median age (range) - years | 45 (1-87) | 36 (1-75) | 45 (2-79) | 48 (28-84) | 64 (33-87) |
| Age group - no./total no. (%) |  |  |  |  |  |
| 0-19 years | 34/539 (6.3%) | 18/115 (15.7%) | 15/331 (4.5%) | 0/57 (0%) | 0/32 (0%) |
| 20-39 years | 190/539 (35.1%) | 46/115 (40.0%) | 122/331 (36.9%) | 16/57 (28.1%) | 4/32 (12.5%) |
| 40-64 years | 255/539 (47.3%) | 42/115 (36.5%) | 170/331 (51.4%) | 28/57 (49.1%) | 13/32 (40.6%) |
| ≥65 years | 60/539 (11.1%) | 9/115 (7.8%) | 23/331 (7.0%) | 13/57 (22.8%) | 15/32 (46.9%) |
| Travel history to Wuhan/Hubei | 205/539 (38.0%) | 39/115 (33.9%) | 135/331 (40.8%) | 23/57 (40.4%) | 8/32 (25.0%) |

95% CI are defined as the percentiles 2.5 and 97.5 of the distribution. Percentages might not total 100% because of rounding.

**Figure 1.**
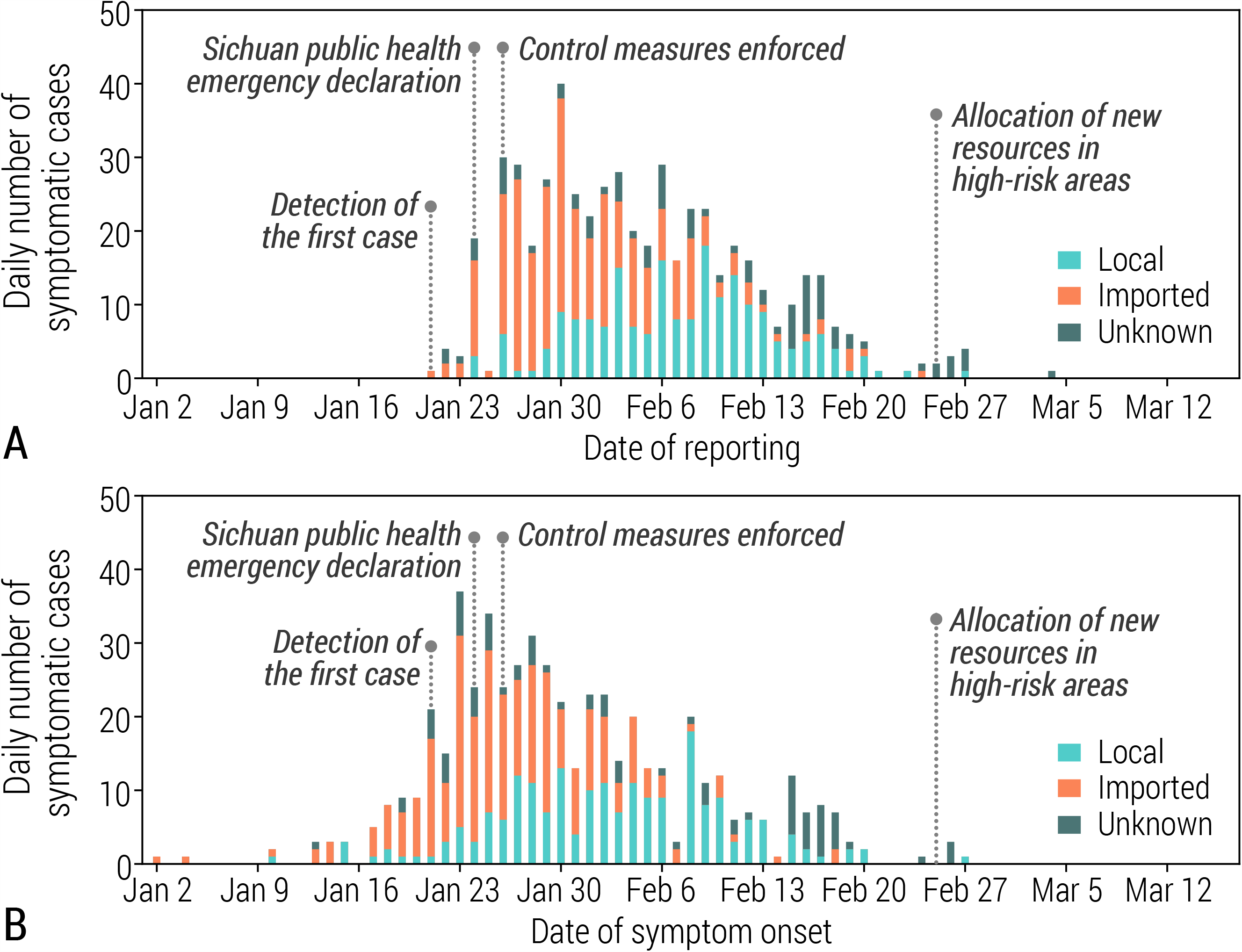
**A** Daily number of new symptomatic cases in Sichuan by date of reporting, disaggregated into cases with travel history to Wuhan/Hubei (imported), cases resulting from local transmission (local), and those for whom the travel history was unknown (unknown). **B** Same as A, but by date of symptom onset.

The epidemic spread undetected in Sichuan Province until January 21, when the first COVID-19 case was identified. In the following days, 44 cases were identified to have symptom onset before that date (Fig. 1b). The first symptomatic locally acquired case reported symptoms on January 10, a few days after the symptom onset of the first two detected cases with travel history from Wuhan. Up to January 30, the epidemic was mostly sustained by imported cases (travel history to or from Wuhan or Hubei), with these peaking around January 23, just before the lockdown was imposed in Wuhan. After that date, we observe a steady decrease in imported cases and the opposite trend in local cases, showing evidence for sustained autochthonous transmission, peaking new local cases in February 8. After February 27, no new symptomatic cases were reported.

The median age of the overall cases was 45 years (1-87) (Tab. 1). This median was higher in local cases (48 years, range: 1-87) than in imported cases (40 years, range: 2-81), see Tab. 2. The severity of cases increased by age (Tab. 1). Overall, the proportion of cases among individuals younger than 18 years was 6.3%, with no severe or critical cases in that age group (Tab. 1).

**Table 2.** Characteristics of laboratory-confirmed COVID-19 cases in Sichuan province disaggregated by locally acquired infections and infected individuals with travel history to Wuhan/Hubei. 95% CI are defined as the percentiles 2.5 and 97.5 of the distribution. Percentages might not total 100% because of rounding.

| Characteristics | Imported cases | Local cases |
| --- | --- | --- |
| Median age (range) - years | 40.0 (2–81) | 48.0 (1–87) |
| Age group - no./total no. (%) |  |  |
| 0-19 years | 17/253 (6.7%) | 12/200 (6.0%) |
| 20-39 years | 103/253 (40.7%) | 58/200 (29.0%) |
| 40-64 years | 120/253 (47.4%) | 99/200 (49.5%) |
| ≥65 years | 13/253 (5.1%) | 31/200 (15.5%) |
| Case severity |  |  |
| Mild | 50 (19.8%) | 46 (23.0%) |
| Symptomatic | 168 (66.4%) | 119 (59.5%) |
| Severe | 25 (9.9%) | 24 (12.0%) |
| Critical | 10 (4.0%) | 10 (5.0%) |
| Male - no./total no. (%) | 144/253 (56.9%) | 97/200 (48.5%) |

Overall, the proportion of male cases was just above 50% for all level of severity, except for mild ones (Tab. 1). This slightly unbalanced proportion of male cases is more evident in imported cases (56.9%, Tab. 2), suggesting a larger fraction of travelers among males.

The mean time interval from symptom onset to hospital admission was estimated at 3.0 days (95%CI: 0.0-14.0, n=153). By considering only cases reported before the declaration of the public health emergency, the mean was estimated at 3.2 days (95%CI:0.0-14.0, n=17) and it decreased to 2.8 days (95% CI: 0.0-14.0, n=136) thereafter. This mean time interval from symptom onset to reporting was estimated at 5.0 days (95%CI: 0.0-17.0, n=539) and followed a decreasing trend over time, from 5.4 days (95%CI:0-18, n=27) before the declaration of the public health emergency to 4.9 days (95%CI:0-16, n=512) thereafter.

### Reproduction number

Led by the first few imported cases from Wuhan/Hubei, we estimated the daily reproductive number to be well below the epidemic threshold at the beginning of the outbreak in Sichuan (Fig. 2). The mean basic reproduction number R_0_ was estimated at 2.4 (95% CI 1.6–3.7) over the period from January 18 to January 24. This figure becomes 2.1 (95% CI 1.6–2.7) if we consider the 2-week period from January 11 to January 24.

**Figure 2.**
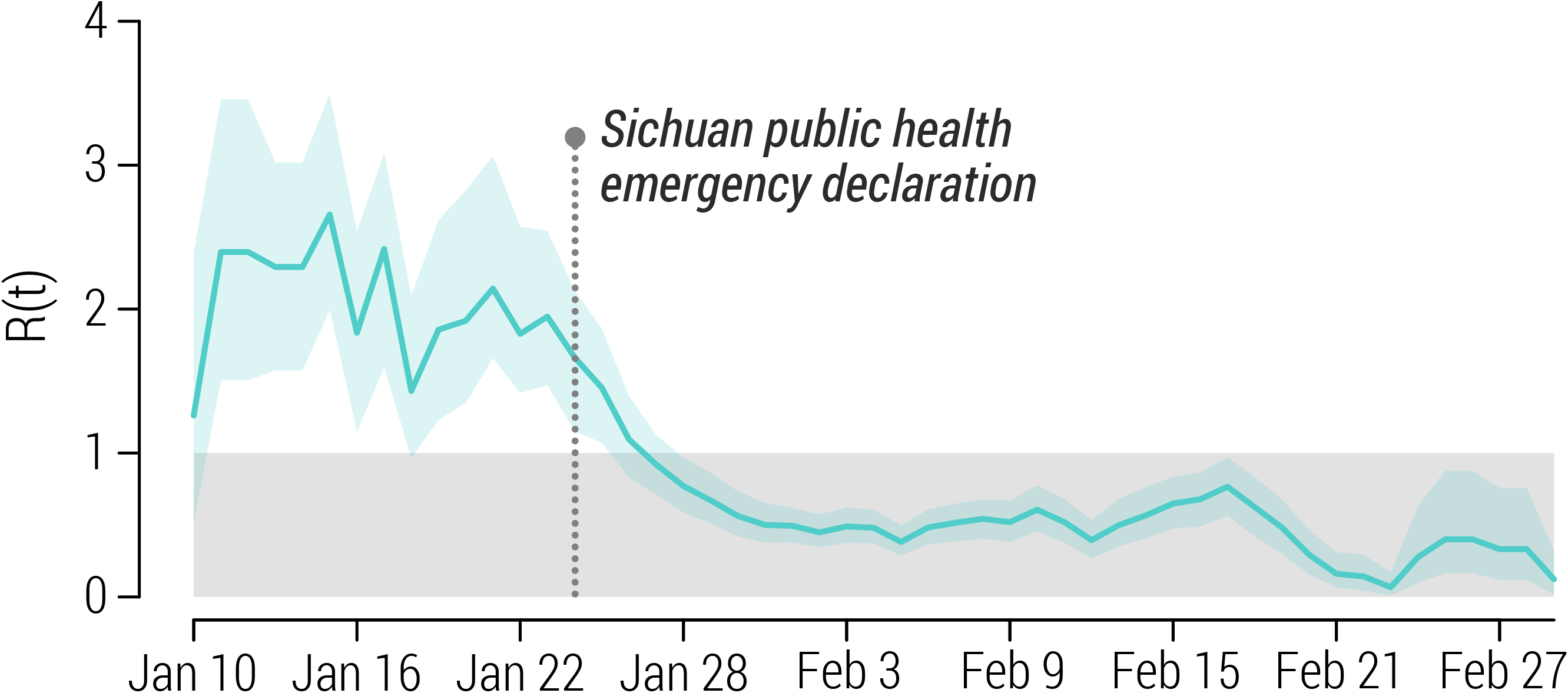
Estimated daily reproduction R(t) (mean and 95% CI) over a 5-day moving average.

We estimated that the epidemic was self-sustained with the mean R(t) above the epidemic threshold for about 2.5 weeks from January 10 to 27. From the declaration of the public health emergency in Sichuan, the estimated R(t) continued to decline with a mean crossing the epidemic threshold on January 27, 3 days after the declaration. Since then, R(t) was estimated to fluctuate constantly below the epidemic threshold (Fig. 2).

### Averted cases

Should the public health emergency been declared one week later, we estimated that the epidemic would have lasted about one week longer. However, should the declaration been done our weeks later, we estimated a non-linear effect, with an epidemic lasting 49 days longer (95%CI: 31-68 days) and the last case reported on April 19 (95%CI April 1-May 10) (Fig. 3).

**Figure 3.**
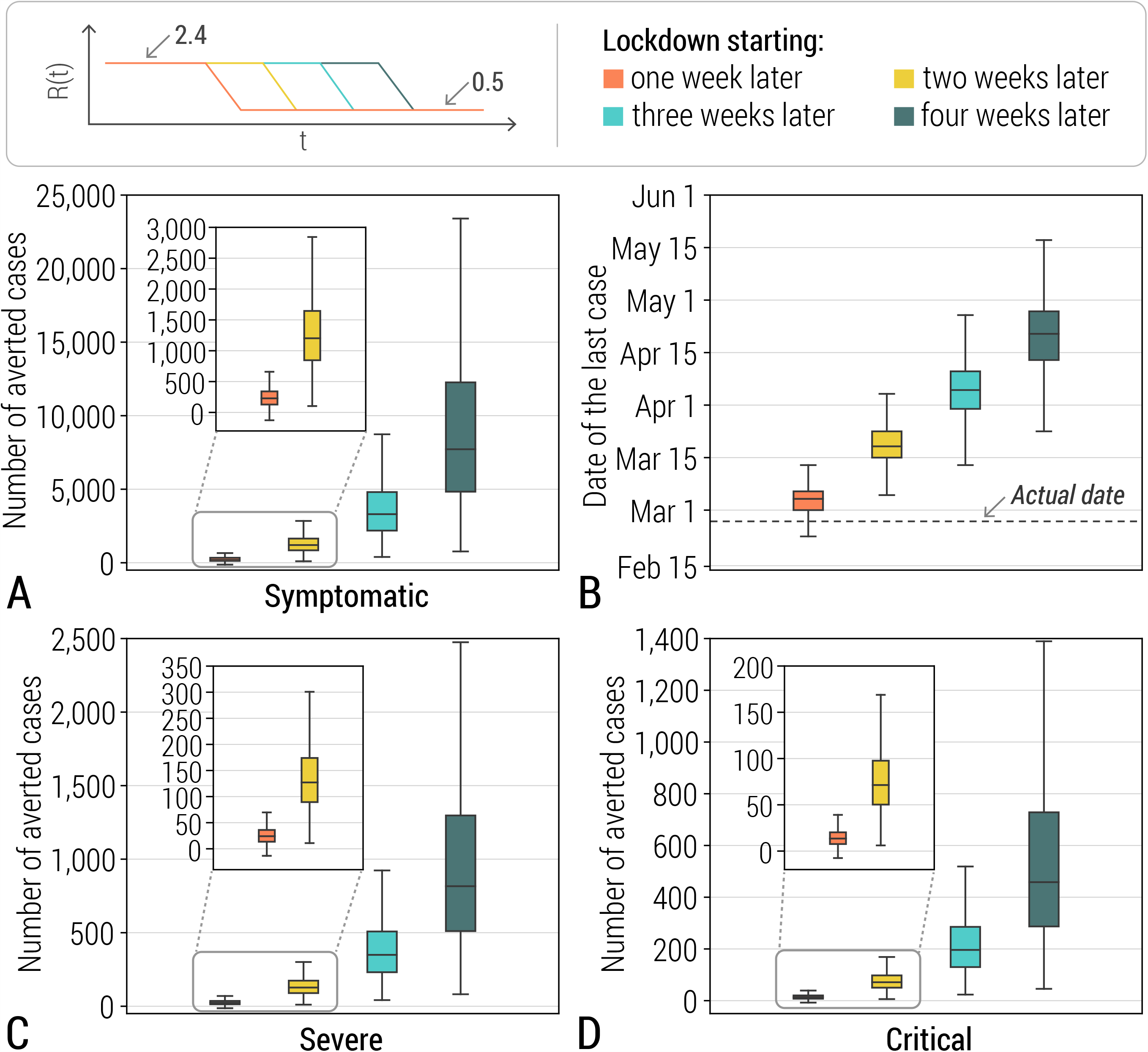
**A** Estimated number of averted cases (min, quantile 0.25, mean, quantile 0.75, max), should the public health declaration have occurred one to four weeks later. Estimates are obtained by considering R_0_=2.4 (95% CI 1.6– 3.7) and R_final_=0.47 (95% 0.4-0.54); R(t) is assumed to follow a 1-week linear decrease from R_0_ to R_final_. R_final_ was estimated over the period from February 1 (i.e., one week after the declaration of the emergency) to the end of the outbreak. **B** Same as A, but for the date of the last case of the simulated epidemics. **C** Same as A, but for severe cases. **D** Same as B, but for critical cases.

The final mean number of projected cases was estimated to be between 775 (95%CI 478-1,113) in the scenario where interventions started one week later than the actual date and 9,755 (95%CI 1,856-26,084) in the scenario considering a 4-week delay. The estimated mean number of averted cases ranged from 236 (95%CI-61-574), including 24 (95%CI-6-60) severe and 14 (95%CI-3-34) critical cases to 9,216 (95%CI 1,317-25,545) cases, including 974 (95%CI 139-2701) severe cases, 547 (95%CI 78-1516) critical cases (Fig. 3).

## 4. DISCUSSION

We provided a characterization of the COVID-19 epidemiology in Sichuan Province of China. The outbreak accounted for a total of 539 PCR positive subjects and was characterized by a combination of local transmission and case importations. We estimated that SARS-CoV-2 transmissibility was above the epidemic threshold for about 4 weeks and then quickly declined after the declaration of a public health emergency in the province and the implementation of strict control measures. We found clear positive effects of the interventions implemented in Sichuan, possibly in combination with an increased awareness of the population about the epidemic spread, which achieved the interruption of transmission leading to a dramatic reduction of the COVID-19 burden in Sichuan.

With a total of 539 cases, Sichuan was among the provinces that were able to successfully contain the COVID-19 outbreak. The epidemic started thanks to the importation of cases from Wuhan/Hubei. Then, we found a clear relationship between the closing of Wuhan province, with only a handful of cases were imported from Wuhan/Hubei since early February. We observe a disproportionate fraction of COVID-19 cases being male in imported cases. In the local cases however we see the reverse. This result indicates potential differential exposure by sex occurring at the beginning of the epidemic and is in overall in line with [13,14], where no significant difference in the risk of infection by gender was found. Nonetheless, it is important to stress that there are several limitations to our characterization of the COVID-19 outbreak in Sichuan that are related to investigation of rapidly evolving novel epidemics, such as biases in the detection of the first few cases, reporting rate, and unknown specifics of a novel pathogen.

In agreement with previous studies focusing on COVID-19 spread in China [10,11,13,14], we found a disproportionately low fraction of cases among individuals younger than 18 years as compared to the age structure of the population. From the data available here, it is not possible to ascertain whether younger individuals have a reduced risk of infection or an increased propensity for a milder clinical outcome of infection (thus resulting in a lower rate of detection). Both hypotheses were already discussed in [15] and found support in empirical epidemiological studies [16,17]. Nonetheless, it has be taken into account that for the most of the epidemic, schools have been closed, not only due to the new year celebrations, but the fact that as the epidemic progressed they did not reopen [3]. Zhang and colleagues [16] showed that children recorded the largest number of contacts among all age groups on a regular weekday due to contacts at schools pre COVID-19, but this was largely reduced after the closure. It is uncertain if nationwide school holidays affected the small proportion of symptomatic cases among school-age individuals. Our approach of documenting the proportion of cases by age and severity does not allow us to readily tease apart the relative roles of age in susceptibility but leaves open opportunity for further scrutiny regarding potential drivers or risk factors that might lead to cohort effects of susceptibility to severe disease.

We estimated that the time interval from symptom onset to hospital admission decreased after the declaration of a public health emergency in Sichuan, suggesting a possible effect of the improved population awareness of the population. Also, the time interval from symptom onset to reporting decreased over time, suggesting an increased effort in case identification and testing from the local health authorities. Both these findings are consistent with those reported in [10]. Nonetheless, although the estimated decrease over time in these two key time intervals may have contributed to the successful control of the COVID-19 outbreak in Sichuan, from the data we have, we cannot provide evidence of their effect.

In the early phase of the outbreak, the daily reproduction number was essentially led by the first few imported cases from Wuhan/Hubei and we estimated the basic reproduction number to be in the ballpark of previous studies about COVID-19 spread in China [11,18-21]. We estimated that the daily reproduction number have followed a decreasing pattern before the declaration of the public health emergency by Sichuan authorities. This may be possible due to the increased awareness of the population of the ongoing COVID-19 epidemic in Wuhan and other areas of China. In fact, it has become apparent that individual and collective human behavior has had subtle and recognizable effects on SARS-CoV-2 transmission [22–25]. After the declaration of the public health emergency on January 24 and the adoption of strict control measures, we estimated a quick reduction of daily reproduction number, similar to what observed in other provinces of China [10] and elsewhere [26-29]. It is important to stress that the case definition changed over the course of the epidemic and, in particular, it was broadened on January 27 to include milder cases [30]. As such, it is possible that we are slightly overestimating the daily reproduction number since then and thus underestimating the beneficial effect of the interventions and population awareness in lowering SARS-CoV-2 transmission potential. However, as shown in [10], the effect is probably not very noticeable.

We found that the implemented control strategies and population awareness have been highly effective in preventing the healthcare system from being overwhelmed by severe and critical COVID-19 cases. In particular, should the health authorities waited four weeks longer to declare the public health emergency, the epidemic would lasted more than six weeks longer and the number of cases would have been of the order of 10,000. It is important to stress that this figure would be much larger if we consider the number of infections instead of cases. In fact, asymptomatic individuals represent a sizable share of SARS-CoV-2 infected individuals [31,32] – up to 73.9% of the infected individuals aged less than 60 years developed neither fever nor respiratory symptoms according to Poletti and colleagues [17]. The evaluation of alternative strategies better targeting asymptomatic individuals is beyond the scope of this work.

It is important to stress that, to estimate the number of averted cases, we adopted the renewal equation to project the final number of cases. Thus we assumed that, the epidemic had continued its spread with the same basic reproduction number estimated during the initial exponential growth phase of the epidemic, before the outbreak was detected. It is however possible that, even if the public health emergency were not timely declared in Sichuan, the population could have adopted significantly different behaviors based on the knowledge that the epidemic was spreading in other areas of China. Moreover, the renewal equation approach is extremely simple and does not account for all the details of the mechanisms of SARS-CoV-2 transmission, such as the influx of imported cases or the underlying structure of the contact network of the population. In particular, we assumed that the distribution of cases by severity would have remained unchanged with respect to that estimated in the early phase of the outbreak. This may have not been the case had the age-distribution of cases changed over time or had healthcare system been overwhelmed. Moreover, we did not account for the depletion of susceptible individuals in the population. However, since we projected cases in a short time window (a few weeks), the depletion of susceptible individuals is negligible and we do not expect to observe dramatic changes in the age distribution of cases over such a short time frame.

## 5. CONCLUSIONS

Our results show the success in control strategies and adaptive behavioral changes of the population were instrumental in interrupting the SARS-CoV-2 transmission in Sichuan Province and preserved the healthcare system from a possible disruptive failure due to overwhelming stress imposed by the large number of severe and critical COVID-19 cases. Nevertheless, it is important to remark that the COVID-19 pandemic is far from being controlled, as we are still far from herd immunity and large proportion of the population remains susceptible. Thus, the course of the pandemic will rely on the efficiency of control strategies and individual behavior in the foreseeable future.

## Data Availability

Data and code will be made available on Figshare upon acceptance of the manuscript.

## Data Accessibility

Data and code will be made available on Figshare upon acceptance of the manuscript.

## Authors’ contributions

Q.-H.L., A.V., and M.A. designed the experiments. K.Y., H.Z., X.Y., and W.Z. collected the data. Q.H.-L., K.Y., and M.A. conducted the analysis. Q.-H.L., S.M., A.V., J.L., H.Y., W.Z., A.I.B., T.Z, and M.A. interpreted the results. A.I.B. and M.A. wrote the manuscript. Q.H.-L., A.V., H.Y., W.Z., and T.Z. edited the manuscript.

## Competing interests

A.V. has received funding from Metabiota. H.Y. has received research funding from Sanofi Pasteur, GlaxoSmithKline, Yichang HEC Changjiang Pharmaceutical Company, and Shanghai Roche Pharmaceutical Company. All other authors declare no competing interests.

## Acknowledgements

The authors thank the West China Biomedical Big Data Center providing the data and the access to the platform for simulations and Nicole Samay for her assistance in preparing the figures.

## Funding

This work was supported by the Special Funds for Prevention and Control of COVID-19 of Sichuan University (0082604151026), Chengdu Science and Technology Bureau (2020-YF05-00073-SN), the Fundamental Research Funds for the Central Universities (1082204112289), the Science and Technology Department of Sichuan Province (2020YFS0007, 2020YFS0009), and the National Natural Science Foundation of China (11975071, 61673085).

## List of Supplementary Figures

**Figure S1.**
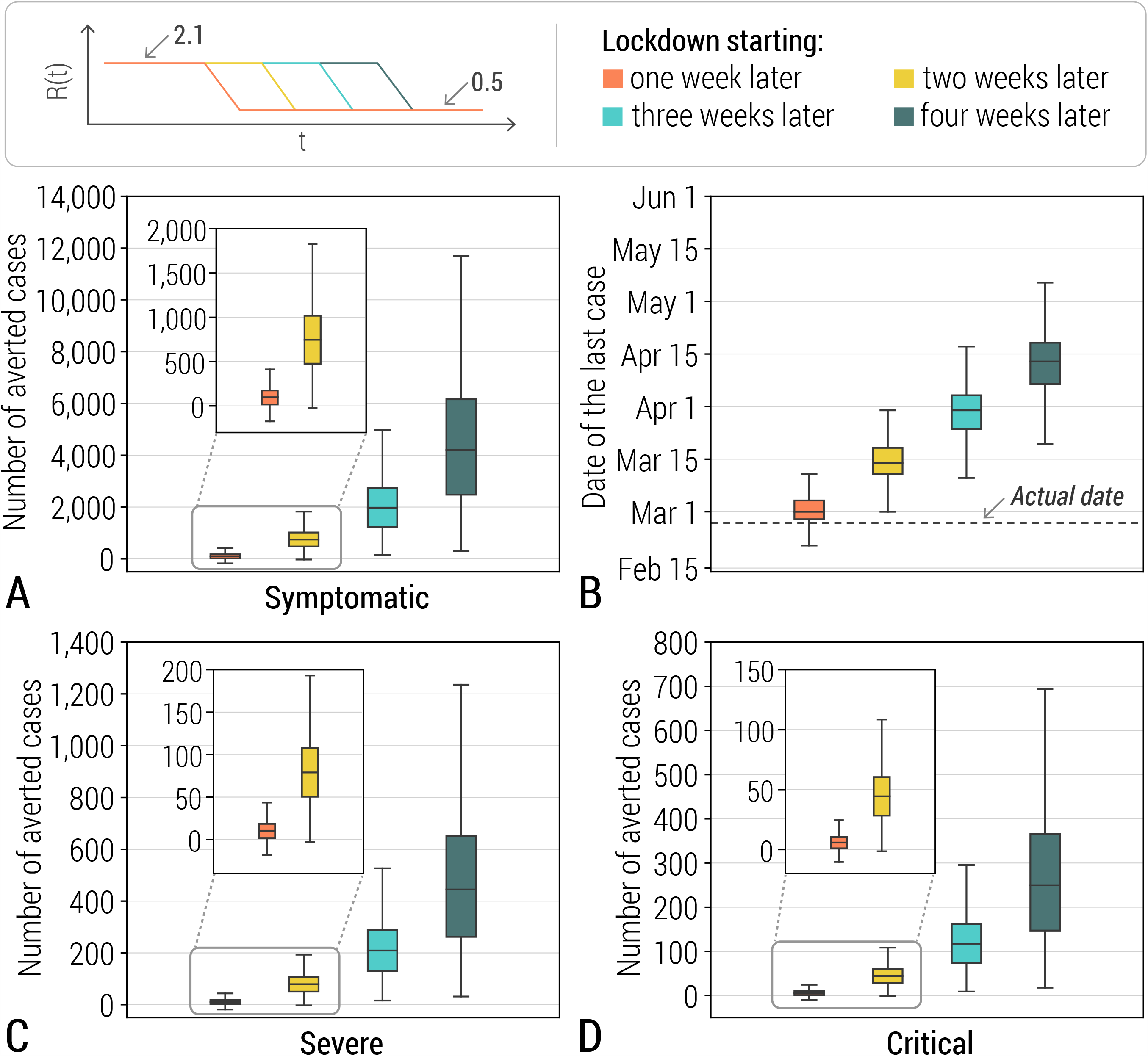
**A** Estimated number of averted cases (min, quantile 0.25, mean, quantile 0.75, max), should the public health declaration have occurred one to four weeks later. Estimates are obtained by considering R_0_=2.1 (95% CI 1.6– 2.7) and R_final_=0.47 (95% 0.4-0.54); R(t) is assumed to follow a 1-week linear decrease from R_0_ to R_final_. R_final_ was estimated over the period from February 1 (i.e., one week after the declaration of the emergency) to the end of the outbreak. **B** Same as A, but for the date of the last case of the simulated epidemics. **C** Same as A, but for severe cases. **D** Same as B, but for critical cases.

**Figure S2.**
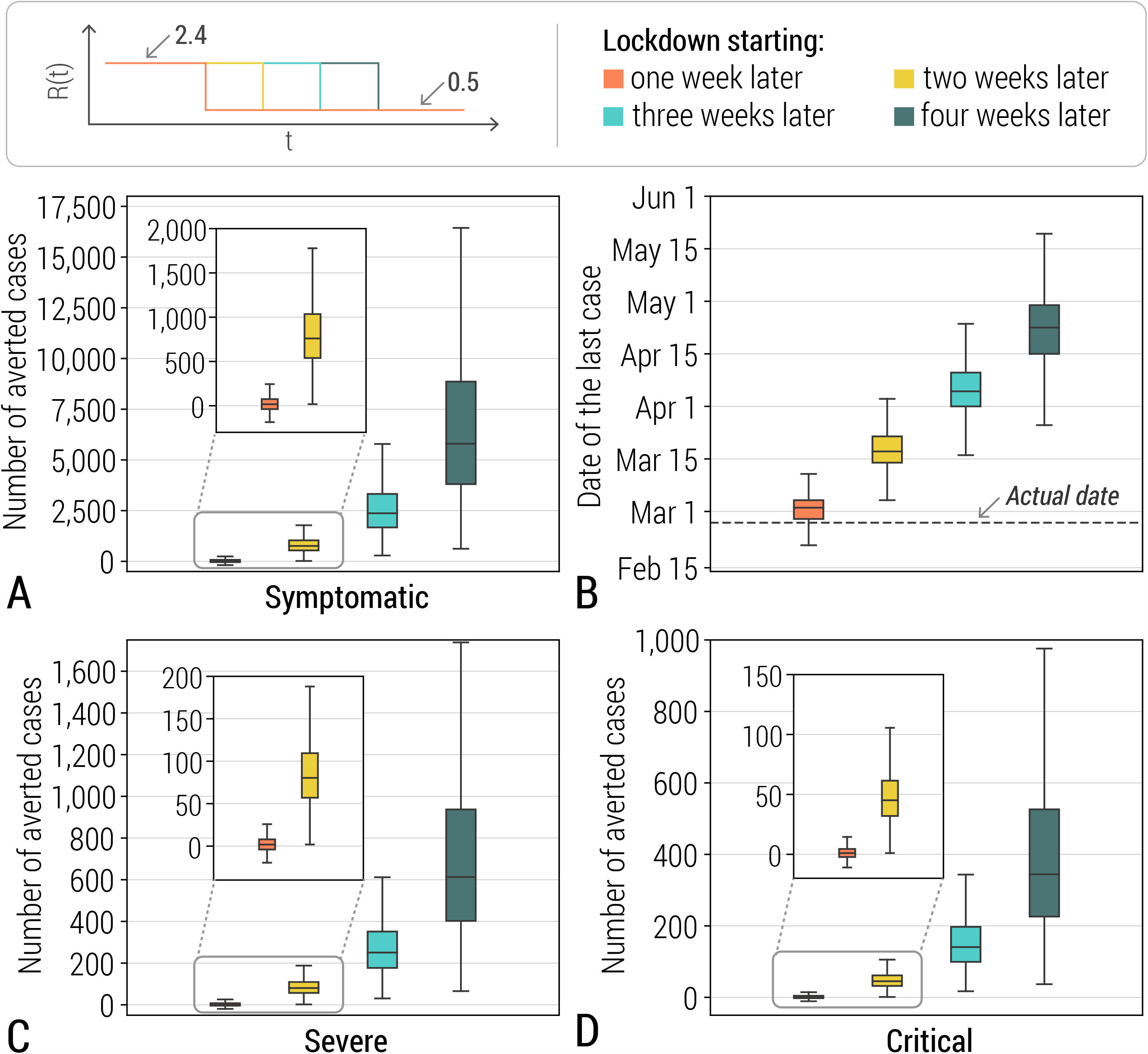
**A** Estimated number of averted cases (min, quantile 0.25, mean, quantile 0.75, max), should the public health declaration have occurred one to four weeks later. Estimates are obtained by considering R_0_=2.4 (95% CI 1.6– 3.7) and R_final=_0.53 (95%CI: 0.47-0.60); R(t) is assumed to instantaneously drop from R_0_ to R_final_. R_final_ was estimated over the period from January 25 (i.e., the day after the declaration of the emergency) to the end of the outbreak. **B** Same as A, but for the date of the last case of the simulated epidemics. **C** Same as A, but for severe cases. **D** Same as B, but for critical cases.

## Notes

### Competing Interest Statement

The authors have declared no competing interest.

